# Attitudes and Perceptions of Biomedical Journal Editors-in-Chief Towards the Use of Artificial Intelligence Chatbots in the Scholarly Publishing Process: A Cross-Sectional Survey

**DOI:** 10.1101/2025.05.26.25328101

**Authors:** Jeremy Y. Ng, Malvika Krishnamurthy, Gursimran Deol, Wid Al-Zahraa Al-Khafaji, Vetrivel Balaji, Magdalene Abebe, Jyot Adhvaryu, Tejas Karrthik, Pranavee Mohanakanthan, Adharva Vellaparambil, Lex M. Bouter, R. Brian Haynes, Alfonso Iorio, Cynthia Lokker, Hervé Maisonneuve, Ana Marušić, David Moher

## Abstract

**Background:** Artificial intelligence chatbots (AICs) are designed to mimic human conversations through text or speech, offering both opportunities and challenges in scholarly publishing. While journal policies of AICs are becoming more defined, there is still a limited understanding of how Editors-in-Chief (EiCs) of biomedical journals’ view these tools. This survey examined EiCs’ attitudes and perceptions, highlighting positive aspects, such as language and grammar support, and concerns regarding setup time, training requirements, and ethical considerations towards the use of AICs in the scholarly publishing process.

**Methods:** A cross-sectional survey was conducted, targeting EiCs of biomedical journals across multiple publishers. Of 3725 journals screened, 3381 eligible emails were identified through web scraping and manual verification. Survey invitations were sent to all identified EiCs. The survey remained open for 5 weeks, with three follow-up email reminders.

**Results:** The survey had a response rate of 16.5% (510 total responses) and a completion rate of 87.0%. Most respondents were familiar with AI chatbots (66.7%), however, most had not utilized chatbots in their editorial work (83.7%) and many expressed interest in further training (64.4%). EiCs acknowledged benefits such as language and grammar support (70.8%) but expressed mixed attitudes on AIC roles in accelerating peer review. Perceptions included the initial time and resources required for setup (83.7%), training needs (83.9%), and ethical considerations (80.6%).

**Conclusions:** This study found that EiCs have mixed attitudes toward AICs, with some EICs acknowledging their potential to enhance editorial efficiency, particularly in tasks like language editing, while others expressed concerns about the ethical implications, the time and resources required for implementation, and the need for additional training.

## INTRODUCTION

Artificial intelligence (AI) systems enable computers or a computer-controlled robot to execute tasks traditionally associated with human cognitive functioning; this includes comprehending complex information, solving problems, and learning from experience.^1–3^ Among these systems, artificial intelligence chatbots (AICs) are AI tools that leverage deep learning AI algorithms to mimic human-like conversation.^4–6^ Popular examples of AICs include ChatGPT, Google Gemini, Meta AI, and Microsoft Co-Pilot.^7–10^

As AICs become increasingly integrated into the academic landscape, it is essential to understand how different stakeholders perceive and regulate their use. Several studies have explored attitudes toward AICs, as well as policies governing their application in scientific research and publishing. A study by Ng et al. investigated the attitudes and perceptions of medical researchers toward AICs in the scientific process through an international cross-sectional survey.^11^ Their findings revealed a notable interest in leveraging AICs for research-related tasks but also highlighted concerns regarding the transparency and accuracy of these tools, which could impact scientific credibility. Bhavsar et al. audited 162 academic publishers and found that only 34.6% had explicit AIC policies.^12^ While most required AI-generated content disclosure while prohibiting AICs as authors, some banned AI-assisted writing entirely.^12^ Their systematic review highlighted the need for clearer, standardized guidelines.^12^ Yin et al. further explored AI policies in medical journals, categorizing guidelines based on scope and enforcement.^13^ These findings shed light on the usage guidelines of generative AI (GAI) across medical journals, emphasizing the need for consistent recommendations. These studies collectively highlight both the opportunities and challenges of AIC integration, underscoring the importance of ongoing discussions and policy refinement for responsible use. AI tools are increasingly being explored to assist in editorial processes such as manuscript submission, peer review, ethical compliance checks, and final decision-making.^14^

Despite these potential advantages, the integration of AICs into the scholarly publishing is still in its early stages and challenges remain. Concerns include the reliability of AICs in detecting AI-generated content and risks to manuscript confidentiality.^15–17^ To mitigate these risks, the International Association of Scientific, Technical & Medical Publishers (STM) established guidelines emphasizing transparency, protection of intellectual property, confidentiality, and ethical integrity in the use of AI tools to ensure that human oversight remains central to editorial decision-making.^18,19^

Studies have also highlighted the potential of AI to streamline manuscript evaluation and enhance editorial workflows.^20–22^ Editors play a pivotal role in managing these workflows, ensuring the quality and integrity of published content.^23^ AI tools have been introduced to assist editors in tasks such as assessing submission quality, verifying completeness, and flagging issues that require further editorial attention.^14^ Among editorial roles, editors-in-chief (EiCs) are particularly influential, responsible for maintaining journal standards, making final publication decisions, and overseeing editorial workflows.^24^ AICs have shown to support EiCs by reducing time spent on manuscript evaluation and improving decision-making efficiency.^25^

Considering the rapid integration into academic editing practices, understanding the perceptions of key stakeholders, is crucial. While previous studies have examined researcher perspectives and publisher policies, few have directly investigated the views of EiCs, those who hold final authority over manuscript decisions and editorial standards.^11^ EiCs are uniquely positioned to shape the ethical, practical, and procedural integration of AICs into editorial workflows. By focusing on this underexplored group, our study provides timely insights into how AICs are being perceived at the highest level of editorial oversight, contributing to a more nuanced understanding of their current and potential role in scholarly publishing.

## METHODS

### Open Science Statement

The protocol for the study was registered on the Open Science Framework (OSF).^26^ All study materials and data can be found on https://doi.org/10.17605/OSF.IO/D7NV5.27 All raw survey data, including open-ended responses with assigned codes, are available in our data repository at https://osf.io/rhqbx.^28^

### Ethical Considerations

Ethics approval was obtained from the Ottawa Hospital Research Institute Research Ethics Board (REB #: 20240334-01H) and informed consent was obtained from all participants before initiating this study. Respondents identifying information, including IP addresses, was not collected to maintain confidentiality.

### Approach and Study Design

An anonymous, cross-sectional, closed survey of EiCs of biomedical journals was conducted to investigate their attitudes and perceptions towards the use of AICs in the scholarly publishing process. As this is a closed survey, only invited EiCs were eligible to participate, and they were instructed not to forward the survey to colleagues, including other EiCs within their network. Following the initial protocol, additional authors were recruited to assist with data collection and analysis. These authors were not involved in the development of the protocol, which accounts for the increase in the number of authors in the final manuscript compared to the original protocol. Due to confidentiality and privacy considerations, the email list of EiCs will not be shared to prevent potential spam or misuse.

### Sampling Framework

A search was conducted for EiCs of biomedical journals across the five largest scholarly publishers by journal count.^29^ A table detailing included journal sources can be found in the Supplementary file, **eTable 1**. These publishers were chosen to ensure a large sample size and diverse representation across biomedical disciplines. The publishers included Springer & BMC (part of Springer Nature), Taylor & Francis, Elsevier, Wiley, and SAGE. For each journal, the journal name, journal URL, EiC name(s), and EiC email(s) were recorded by a custom web scraping software, which automated journal listing navigation and implemented a prediction algorithm to assess email accuracy. Further details on the web scraping methodology, including software implementation and data verification, are provided in the Web Scraping Methodology section.^30^ While open source, an RRID is not currently available.^30^ The information web scraped was manually verified by a team of research assistants (MK, WAA, MA, JA, TK, PM, and AV) at each stage to remove any potential errors or duplicates before survey administration. When EiC emails could not be found by the software, our team conducted reverse searches using the EiC’s name to find the most recent EiC email on relevant websites including PubMed, faculty webpages, institutional websites, and publisher-specific pages.

### Web Scraping Methodology

A custom, open source, software identified and collected the email addresses of EiCs associated with the academic journal categories.^30^ The software was developed in Python 3.9 and employed libraries such as BeautifulSoup4 for parsing HTML content, Requests for making HTTP requests, Pandas for data manipulation and export, tqdm for progress tracking, and Selenium WebDriver for automated browser navigation when needed. The software collected EiC names from each journal and accounted for instances of multiple EiCs per journal by specifically searching for HTML elements tagged in a manner that denotes the EiC(s) information. PubMed was then queried for articles authored by the identified EiCs to collect the email addresses. The software extracted email addresses found within the affiliation or contact information sections. Given the potential for multiple email addresses, the script employed a prediction algorithm.^30^ This algorithm compared each extracted email address against the EiC’s name, assigning a score based on likelihood of accuracy, and prioritized addresses that appeared more personalized (e.g., containing parts of the EiC’s name).^30^ If no email addresses met the accuracy criterion, the script recorded this outcome, ensuring transparency in data collection. All screened journals were compiled into a structured dataset, including EiC name, journal name, journal URL, extracted email addresses, and accuracy score.^31^ This dataset can be found in our data repository at https://osf.io/dw9rs with EiC names and email addresses excluded to maintain confidentiality and prevent susceptibility to spam.^31^

### Eligibility Criteria

Participants must have self-identified as an EiC of a scholarly journal of any biomedical topic (e.g., medicine/surgery, dentistry, nursing, pharmacy, and other health-related topics and professions), and could have held the role of EiC for any length of time. Participants who were not an EiC of a scholarly journal on any biomedical topic were ineligible.

### Participant Recruitment

Convenience sampling was used to recruit participants. An invitation authorized by our local REB detailing the study and its objectives, was sent to participants through email using Microsoft Outlook Mail Merge software.^32^ The invitation can be found in Supplementary File, eMethods. When participants clicked on the survey link, the first page of the survey was an informed consent form where participants had to click “Next” to proceed.

After each round of emails were sent, 16 incorrect emails were either replaced with the correct one or a search was conducted for the correct email in the following situations: 1) if a reply or autoreply was received indicating that the email address was no longer active (e.g., the EiC has changed institutions and had an updated email address), 2) if a response from the invitee indicated that they were not the EiC (e.g., the EiC of the journal had recently changed), or 3) an email was erroneously sent to an individual who has the same name as the EiC. Participation was entirely voluntary, with the option to withdraw at any point prior to submission of the survey. Monetary compensation was not provided.

### Survey

The survey began with a screening question to determine eligibility, followed by questions about demographic information, experience with AICs, the role of AICs in publishing, and the perceived benefits and challenges of AICs. Follow-up questions were included, and a final open-ended question allowed for additional feedback. The survey consisted of 31 questions across seven pages and took about 15 minutes to complete, with participants able to review and modify their answers before submission. The complete survey is provided on OSF: https://osf.io/mzagw.^33^ The first draft of the survey was designed by JYN and reviewed by the co-authors. The survey was pilot-tested prior to distribution by a team of researchers, and their feedback was integrated into the survey.

The survey was administered online through the University of Ottawa’s version of the SurveyMonkey software.^34^ The survey was open for 5 weeks, from July 22, 2024, to August 25, 2024, with three follow-up email reminders sent during this period. EiCs who had not responded to the original invitation received follow-up emails during the weeks of July 29, 2024, August 5, 2024, and August 12, 2024. A 2-week waiting period followed the final reminder.

### Data Management & Analysis

Data collected in the survey was summarized using descriptive statistics, including percentages and frequencies. Stratification across variables (e.g., age, sex, career stage) was employed to determine potential differences in attitudes and perceptions between subgroups of participants. SurveyMonkey’s cross-tabulation feature automatically segmented responses and generated contingency tables, displaying response distributions across groups.^35^ The survey dataset was exported and analyzed using Microsoft Excel. Strengthening the Reporting of Observational Studies in Epidemiology (STROBE) and Checklist for Reporting Results of Internet E-Surveys (CHERRIES) guidelines were used to inform the reporting of this survey study.^36,37^

Qualitative data gathered from open-ended questions was analyzed using thematic content analysis.^38–40^ Pilot coding was conducted by two authors (MK and GD), independently coding all responses to question 14, the first open-ended question in the survey. Based on the outcomes of this independent coding, the authors collaboratively developed a shared codebook through discussion and consensus. Once the codebook was finalized, all remaining open-ended responses were independently coded by the same two authors using the agreed-upon codes and finalized through consensus. Following this, the codes were iteratively refined, and any discrepancies were resolved through discussion. The initial codes were then grouped into overarching themes, based on patterns, similarities, and commonalities observed in the data.^38,39^ These themes were first developed independently by MK and GD and then finalized through collaborative discussion. A descriptive definition for each theme was created to ensure clarity and consistency in interpretation.

## RESULTS

### Respondent Demographics

A total of 3381 survey invitations were sent, and 295 emails bounced (i.e., survey invitation did not reach the participant). From the total emails sent, the publisher distributions for participating EiCs are as follows: 1107 Elsevier, 828 Springer Nature, 730 Taylor and Francis, 464 Wiley, and 252 SAGE. 510 invitees provided responses (17% response rate, n=510/3086), wherein 505 respondents met the eligibility criteria (99%, n=505/510), and 439 completed the survey (87% completion rate, n=439/505). Since not all respondents completed all survey questions, the number of responses for each question was variable. Responses were stratified by age, career stage, research category, and sex can be found in our data repository at https://osf.io/3rycf.^41^

Demographic data is presented in **eTable 2**. Most respondents identified as male (68%, n=311/490), around a third identified as female (31%, n=150/490), and 9 respondents preferred not to say (2%, n=9/490). Most respondents fell into the following three age ranges: 65+ (32%, n=156/492), 56-65 (28%, n=139/492), and 46-55 (27%, n=131/492). Respondents self-identified as coming from 43 countries, across six continents, with the top three countries being the United States of America (40%, n=196/486), United Kingdom of Great Britain and Northern Ireland (10%, n=48/486), and Australia (7%, n=35/486). Most respondents were senior researchers (95%, n=470/493), faculty members at a university/academic institution (78%, n=385/492) and primarily received clinical research manuscript submissions (64%, n=313/492).

### Experience with Artificial Intelligence Chatbots

Many respondents (67%, n=325/487) reported that they were familiar with the concept of AICs and mostly used ChatGPT (67%, n=327/485). The majority of EiCs had not used an AIC for a purpose related to their role as EiC (84%, n=401/479), however some reported using ChatGPT (13%, n=63/479), Microsoft Copilot (2%, n=10/479), Google Gemini (1%, n=5/479), and Meta AI (0.2%, n=1/479) for purposes relating to their role as EiC. **eFigure 1** in the Supplementary File details EiC use of various LLMs for any use and EiC-specific use.

**Figure 1:**
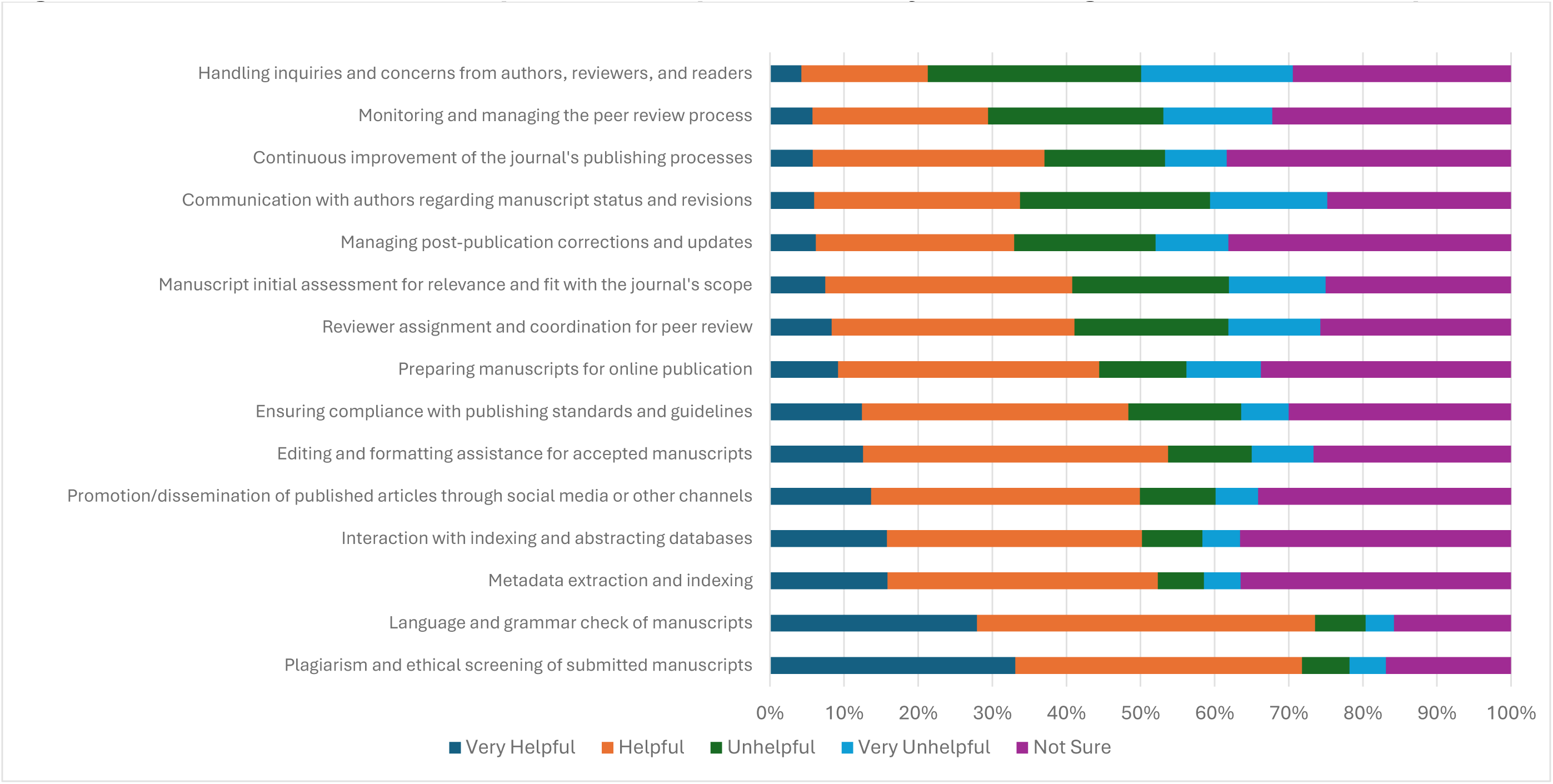
Biomedical EICs Perceptions of Steps in Scholarly Publishing Where AICs Are Helpful.

Respondents also indicated that they were either unlikely (25%, n=123/487) or very unlikely (26%, n=127/487) to use an AIC within the context of their role as an EiC in the future. Most respondents also reported that their journal or publisher either does not provide any training on the use of AICs in context of their role as EiC (54%, n=262/486) or that they were unsure if any training was provided (30%, n=147/486). When asked if the journal or publisher has implemented any policies surrounding the use of AICs in the scholarly publishing process, around half of the respondents answered yes (50%, n=240/484), and the remaining respondents were divided between being unsure of these policies (29%, n=138/484) or indicating that there were no policies implemented (22%, n=106/484). The responses were collected anonymously due to REB restrictions, preventing analysis by publisher.

Respondents highlighted that EiCs should undergo either a lot (45%, n=211/468) or some (44%, n=208/468) training to effectively use AICs in scholarly publishing. They indicated interest in learning more and receiving more training surrounding the use of AICs (64%, n=302/469). e**Table 2** details experience with AICs and their affiliated publisher and journal’s views on policies and training on the use of AICs.

### Role of Artificial Intelligence Chatbots in the Publishing Process

The respondents were asked to rate their agreement with statements on a 5-point scale from “very helpful” to “very unhelpful” regarding how helpful an AIC would be in different steps of the scholarly publishing process (**Figure 1**). Notably, many respondents felt that AICs would be helpful or very helpful in the language and grammar checks (74%, n=345/469) and screening for plagiarism and ethics issues (72%, n=336/468). However, when asked if AICs would assist with the continuous improvement of the journal’s publishing process, although a third believed AICs would be helpful (31%, n=146/467), a notable proportion remained unsure (38%, n=179/467). Similarly, though some respondents believed that AICs would be helpful in managing post-publication corrections and manuscript updates (27%, n=125/467), other respondents were unsure (38%, n=178/467).

Respondents were asked to rate how often they had used AICs in different steps of the scholarly publishing process on a 5-point scale from “Always” to “Never.” (**Figure 2**). The question was answered by 81 respondents and skipped by 429 participants. Most respondents answered that they never have used AICs in any step of the publishing process. Over 80% had never used AICs for monitoring and managing the peer review process (82%, n=65/79), for metadata extraction and indexing (85%, n=68/80), to assist with managing post-publication corrections and updates (86%, n=70/81), interacting with indexing and abstracting databases (83%, n=67/81), or for the continuous improvement of the journal’s publishing process (82%, n=66/81). AICs were often or always used for language and grammar checks of manuscripts (20%, n=17/81) or for plagiarism and ethical screening of the submitted manuscripts (20%, n=17/81). The respondents believed that AICs will be important or very important in the future of scientific research (77%, n=363/470) and that they would have a positive impact on future research outcomes (44%, n=208/471). Similarly, respondents also believed that AICs would be important or very important for improving the scholarly publishing process (79%, n=370/469) and have a positive impact on the future of scholarly publishing (47%, n=220/470). **eFigure 2** in the Supplementary File details EiC perceptions of potential future importance of AICs in scholarly publishing and scientific research, and **eFigure 3** in the Supplementary File details EiC perceptions of potential future impact of AICs in scholarly publishing and scientific research.

**Figure 2:**
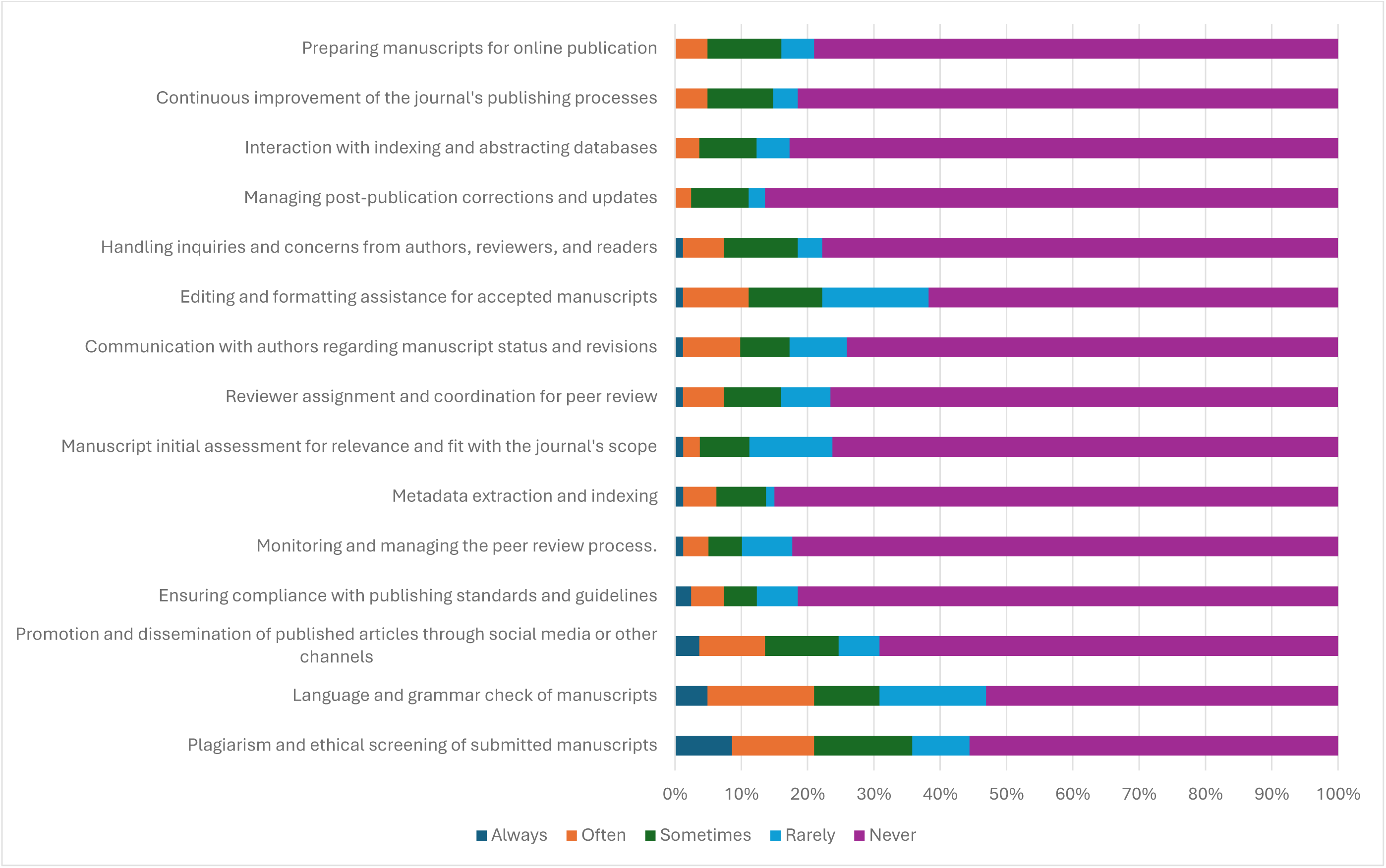
Biomedical EICs Frequency of AIC Use in Various Steps of the Scholarly Publishing Process.

**Figure 3:**
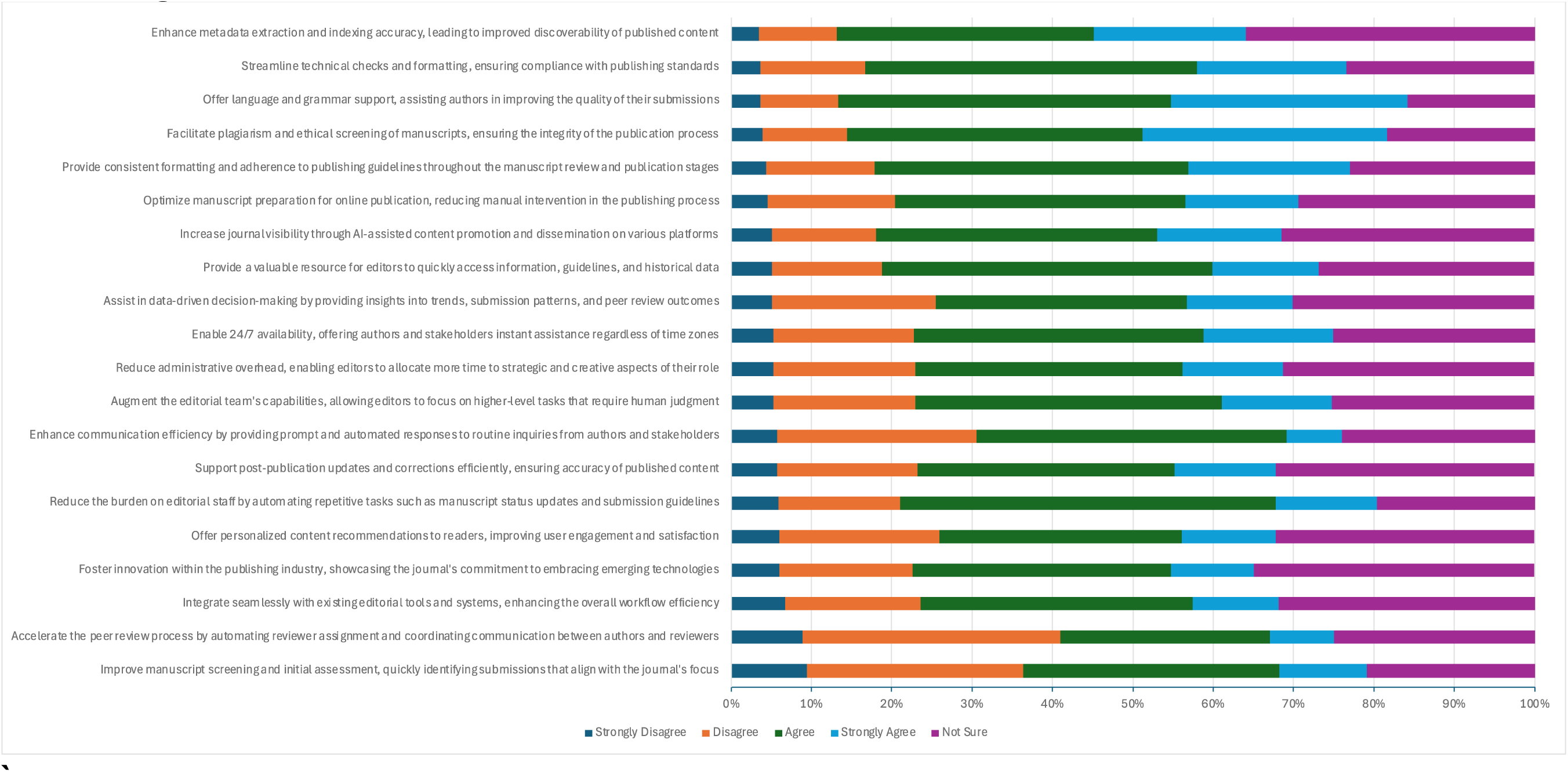
Biomedical EICs Perceptions of Proposed Benefits of AIC Chatbots in the Scholarly Publishing Process.

### Perceived Benefits & Challenges of Artificial Intelligence Chatbots in the Scholarly Publishing Process

The respondents were asked to rate their agreement with statements about some of the proposed benefits to the use of AI chatbots in the scholarly publishing process on a 5-point scale ranging from “Strongly disagree” to “Strongly agree” (**Figure 3**). The respondents agreed and strongly agreed that AICs could reduce the burden on editorial staff by automating repetitive tasks such as manuscript status updates and submission guidelines (59%, n=260/438), offering language and grammar support (71%, n=308/435), facilitating plagiarism and ethics screening of manuscripts (67%, n=294/437), and streamlining technical checks and formatting (60%, n=261/436).

When asked if AICs could accelerate the peer review process by automating reviewer assignment and coordinating communication, ratings were mixed; 179/437 respondents disagreed or strongly disagreed (41%), 149/437 agreed or strongly agreed (34%), and 109/437 respondents were unsure (25%). Similarly, when asked if AICs could help foster innovation within the publishing industry, opinions were also divided; a total of 113/433 respondents disagreed or strongly disagreed (23%), 184/433 agreed or strongly agreed (43%), and 151/437 respondents were unsure (35%).

The respondents were also asked to rate their agreement with statements about some of the potential challenges to the use of AI chatbots in the scholarly publishing process on a 5-point scale ranging from “Strongly disagree” to “Strongly agree.” **Figure 4** depicts the respondents’ answers. The most agreed upon challenges to the use of AICs in the publishing process include: the additional training and monitoring for editors to effectively interact with and oversee AIC interactions (84%, n=360/429), the initial investment of time and resources for setup, customization, and alignment with specific editorial needs (84%, n=359/429), present unforeseen ethical quandaries related to accountability, transparency, and the responsible use of AI technology (81%, n=345/428), risk of biased decisions (80%, n=344/431), struggle to adequately address complex ethical issues that require human judgment and ethical considerations (79%, n=340/429), lack of human insight needed to understand contextual nuances and emotional aspects of interactions with authors and reviewers (78%, n=338/431), susceptibility to technical dependence and failures (75%, n=322/428), and facing user acceptance challenges (72%, n=309/430).

**Figure 4:**
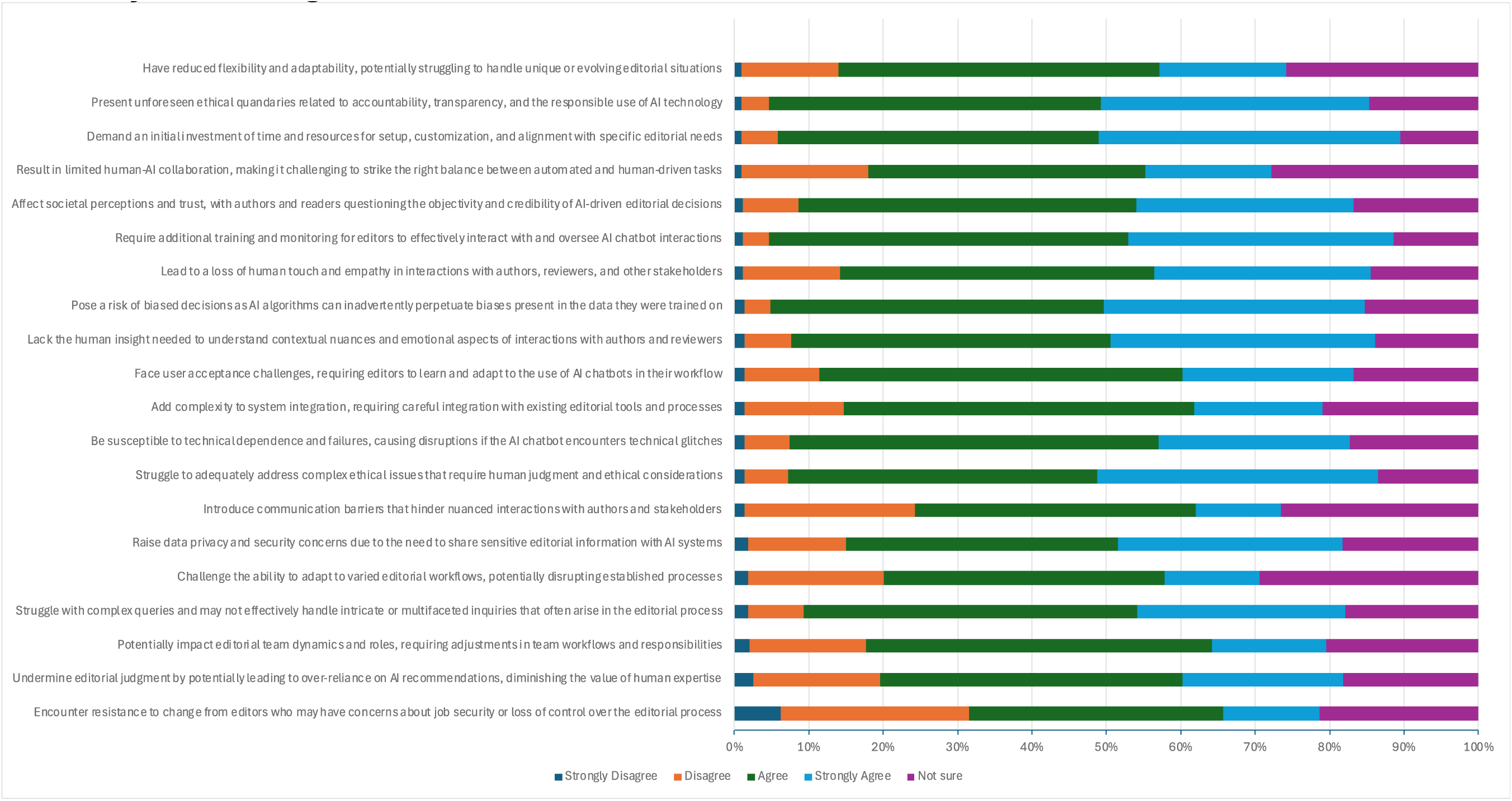
Biomedical EICs Perceptions of Proposed Challenges with AIC Chatbots in the Scholarly Publishing Process.

### Open-Ended Questions

A total of seven optional open-ended questions offered participants an opportunity to share additional insights and feedback on the use of AICs in the scholarly publishing process. The identified themes, subthemes, individual codes, and their frequencies are detailed in supplementary materials available in our data repository at https://osf.io/juzhb.^42^ The main themes identified through the thematic analysis include: “no AI in authorship or peer review” referring to the EiC current journal/publisher policy on AIC use, “ethical, integrity, and privacy concerns” referring to EiC perceptions of challenges with the use of AICs in the scholarly publishing process, and “need for editor AIC training” which refers to need for editor AIC training within the context of the scholarly publishing process. Additional details on these themes are available in the Supplementary File, **eResults** “Themes from Thematic Analysis of Open-Ended Questions).

## DISCUSSION

In this survey, although most respondents were familiar with the concept of AICs, over 80% have never used an AIC for a purpose relating to their role as EiC. Respondents expressed mixed views about the potential use of AICs within their role as EiC, mostly responding that they would be unlikely or very unlikely to do so. While many respondents recognized that AICs could be helpful in tasks like language checks and plagiarism screening, they were generally unlikely to adopt them in their editorial work. Respondents also had mixed views on the benefits of AICs, but there was strong agreement on the potential challenges, including concerns about bias, ethical dilemmas, and technical failures. These findings highlight that, although some benefits of AICs are known, their practical adoption in EiC workflows remains limited.

Respondents generally had a mix of positive and negative views on the use of AICs in scholarly publishing. Many acknowledged that AICs could provide benefits, especially in tasks like language and grammar support (70% of respondents), as well as plagiarism and ethics screening (72%). However, concerns about ethical implications, technical issues, and the resources needed to train and implement these tools led to significant reluctance. A large portion of EiCs expressed the need for further training (64%) and perceived the setup and resource investment as a major barrier to adoption (83%). Despite the limited use of AICs by EiCs, many respondents believed AICs would play an important or very important in the future of scholarly publishing, with 90% believing that some or a lot of training would be required to effectively use AICs in the scholarly publishing process. However, only half reported that their journal or publisher had implemented policies on AIC use, and over 53% indicated that no training was offered. This lack of training and clear policies could be attributed to the relatively new integration of AICs into the publishing process. These findings highlight a need for better tools and training initiatives to ensure the responsible and effective use of AICs. While training is important, it is possible that providing better AI tools and ensuring that editorial staff are proficient with them may also be crucial for EiCs. Moreover, further exploration is required to address ethical considerations, technical challenges, and the potential for misuse in the scholarly publishing landscape. Developing scalable and efficient solutions such as mandatory watermarking to enable detection of AIC outputs, as well as creating and adhering to policies such as the STM guidelines, may help ensure transparency and accountability in the use of AI-generated content.^16,19^

The use of AICs in scholarly publishing is an emerging topic, recognized for its potential to automate and streamline editorial processes. A review by Kousha et al., showed that AICs assisting in initial manuscript evaluations, performing tasks like plagiarism detection and technical checks, reduces editorial workload.^43^ Our study aligns with these findings, where EiCs recognized AICs’ utility in performing routine tasks such as language checks and plagiarism screening. However, similar to concerns highlighted by our respondents, AIC reliability in detecting AI-generated content or fabricated data remains a limitation.^15^ This concern is echoed in our survey and by other studies, such as those by Ng et al. where perceptions towards AICs in both educational and scientific contexts, revealed mixed attitudes regarding their reliability and ethical implications.^11,44^ In educational contexts, Ng et al. investigated the perspectives of biomedical students and postdoctoral fellows on using AICs.^44^ The study revealed that while participants acknowledged the potential of AICs to support learning, they raised significant concerns about academic integrity and the ethical implications of relying on such tools in educational settings.^44^ A similar study by Ng et al. examined the perspectives of medical researchers regarding the role of AICs in the scientific process.^11^ This research found notable interest in employing AICs for research support, but also highlighted concerns about the transparency of AICs and the accuracy of their outputs, which could impact the credibility of scientific research.^11^

Fiorillo & Mehta explored AICs’ potential to reduce editorial bias but also noted risks like algorithmic bias and data manipulation.^21^ These concerns resonate with those voiced by EiCs in our study, particularly regarding the transparency and integrity of using AICs in the publishing process. Ethical frameworks, including those from Committee on Publication Ethics (COPE), the World Association of Medical Editors (WAME), and the European Commission emphasize the importance of transparency in the use of AICs and recommend disclosure to authors without explicitly requiring written consent.^45–47^ There is also current development of initiatives such as the “ChatGPT and Artificial Intelligence Natural Large Language Models for Accountable Reporting and Use Guidelines” (CANGARU) which aim to establish standards for the ethical application of large language models (LLMs) such as ChatGPT in scientific research.^48^ As AIC use continues to expand in academic publishing, the benefits must be weighed against challenges such as bias, data security, and peer-review integrity.

Currently, there are notable concerns and challenges with AIC usage that require immediate attention. While most EiCs were familiar with AICs, the majority had never used them in their editing roles. Many also expressed interests in training, but few reported formal training programs or policies at their journals. This highlights the need for structured training to navigate AIC use effectively and ethically. Clear guidelines and policies are crucial to mitigate risks to research integrity as AICs become more integrated. Respondents also raised concerns regarding bias, ethical, and technical limitations associated with AICs. AI engineers and evaluators should also focus on addressing the current limitations and errors in AIC models to improve their reliability and functionality.

The high rate of skipped responses to the question regarding AIC use in specific publishing steps warrants further exploration. This may indicate discomfort or uncertainty among participants, potentially due to limited familiarity, lack of experience with the tool, or hesitancy to disclose practices due to perceived judgment. Future research should investigate these barriers to promote transparency of AIC use in scholarly publishing.

To track the evolution of attitudes and perceptions of biomedical journal EiCs towards the use of AICs in the scholarly publishing process, the survey should be replicated every two to five years, allowing for longitudinal analysis of trends, emerging concerns, and shifts in editorial policies.

### Strengths and Limitations

This study, based on a large, purposefully selected sample of biomedical journal EiCs, offers broad relevance to the field. By collection of most recent EiC contact information through PubMed and institutional webpages, we minimized the risk of sending survey invitations to invalid or inactive email addresses. Additionally, tracking email bounce-backs allowed for an accurate assessment of survey response rates.

A key limitation of this study is the exclusion of non-English speaking EiCs due to language constraints, as well as the likely underrepresentation of editors from low-and middle-income countries (LMICs) and smaller publishers.^49^ Including EiCs from LMICs could provide a broader range of insights, especially as the challenges and experiences with AI and chatbots within these regions may differ from high income countries.^50^ Moreover, EiCs from smaller publishers, such as society-based or university-based publishers, were not specifically searched for, leading to potential underrepresentation of these groups.^51^ The focus on larger publishers may further limit the generalizability of the findings, as perspectives may differ substantially in terms of AIC usage and challenges. Additionally, the low response rate for our survey may reflect concerns about cyber scams, survey fatigue, and/or unfamiliarity or lack of interest with the subject; however, the high completion rate mitigates some concerns about data quality. Respondents’ lack of familiarity with specific AIC models could affect the depth and accuracy of their responses, especially as AICs rapidly evolve; a "snapshot" of AICs’ current use has limited value, as new capabilities and challenges continue to emerge. We must also acknowledge non-response bias and selective response bias, which may affect the internal validity and generalizability of the collected data.^50^ It is important to note that some aspects of AIC use may overlap, particularly regarding editorial tasks and functions such as monitoring and managing the peer review process. While the survey aimed to differentiate distinct stages of the publishing workflow, variation in respondents’ interpretations may have influenced the reported data. While the use of a Likert scale for attitude measurement increases the reliability of the responses, the survey relied on self-reported data, which is susceptible to bias, including social desirability bias, where participants may respond in a way they believe is more socially acceptable.^52^ Additionally, the closed nature of the survey may limit generalizability, as it was restricted to EiCs of journals within specific publishers and did not include broader representation from other scholarly fields.

## CONCLUSION

In conclusion, this study provides valuable insights into the attitudes and perceptions of biomedical journal Editors-in-Chief regarding the use of artificial intelligence chatbots in the scholarly publishing process. While many EiCs acknowledge the potential of AICs to enhance editorial efficiency, particularly in language editing and plagiarism detection, there remains significant hesitation regarding their broader integration due to concerns about ethical implications, training needs, and resource constraints. The findings highlight the need for clearer guidelines, training programs, and ethical frameworks to facilitate the responsible adoption of AICs in scholarly publishing. As AI technology continues to evolve, future research should explore longitudinal trends in editorial perspectives, assess the impact of AI adoption on publication quality and integrity, and develop best practices for integrating AI tools in academic publishing workflows. Addressing these challenges proactively will ensure that AICs serve as a valuable complement to, rather than a replacement for, human editorial expertise.

## Data Availability

All data and materials associated with this study are contained within this manuscript or have been posted on the Open Science Framework.

https://doi.org/10.17605/OSF.IO/D7NV5

## ABBREVIATIONS

AI: artificial intelligence
AIC: artificial intelligence chatbot
CANGARU: ChatGPT and Artificial Intelligence Natural Large Language Models for Accountable Reporting and Use Guidelines
ChatGPT: chat generative pre-trained transformer
COPE: Committee on Publication Ethics
EiC: editor-in-chief
LLM: large language model
LMIC: low- and middle-income countries
OSF: Open Science Framework
REB: research ethics board
STM: International Association of Scientific, Technical & Medical Publishers
STROBE: Strengthening the Reporting of Observational Studies in Epidemiology
WAME: World Association of Medical Editors

## Declarations

### Ethics Approval and Consent to Participate

Ethics approval was granted by the Ottawa Health Science Network Research Ethics Board (REB Number: 20240334-01H).

### Consent for Publication

All authors consent to this manuscript’s publication.

### Availability of Data and Materials

All data and materials associated with this study are contained within this manuscript or have been posted on the Open Science Framework and can be found here: https://doi.org/10.17605/OSF.IO/D7NV5

### Authors’ Contributions

JYN: designed and conceptualized the study, co-drafted the manuscript, and gave final approval of the version to be published.

MK: was the main results drafter, analysed data, co-drafted the manuscript, and gave final approval of the version to be published.

GD: analysed data, co-drafted the manuscript, and gave final approval of the version to be published.

WAA: assisted with data analysis, made critical revisions to the manuscript, and gave final approval of the version to be published.

VB: developed web scraping automation, made critical revisions to the manuscript, and gave final approval of the version to be published.

MA: completed manual verifications of EiC information, made critical revisions to the manuscript, and gave final approval of the version to be published.

JA: completed manual verifications of EiC information, made critical revisions to the manuscript, and gave final approval of the version to be published.

TK: completed manual verifications of EiC information, made critical revisions to the manuscript, and gave final approval of the version to be published.

PM: completed manual verifications of EiC information, made critical revisions to the manuscript, and gave final approval of the version to be published.

AV: completed manual verifications of EiC information, made critical revisions to the manuscript, and gave final approval of the version to be published.

LMB: assisted with the design of the study and the analysis of data, made critical revisions to the manuscript, and gave final approval of the version to be published.

RBH: assisted with the design of the study and the analysis of data, made critical revisions to the manuscript, and gave final approval of the version to be published.

AI: assisted with the design of the study and the analysis of data, made critical revisions to the manuscript, and gave final approval of the version to be published.

CL: assisted with the design of the study and the analysis of data, made critical revisions to the manuscript, and gave final approval of the version to be published.

HM: assisted with the design of the study and the analysis of data, made critical revisions to the manuscript, and gave final approval of the version to be published.

AM: assisted with the design of the study and the analysis of data, made critical revisions to the manuscript, and gave final approval of the version to be published.

DM: assisted with the design of the study and the analysis of data, made critical revisions to the manuscript, and gave final approval of the version to be published.

## Acknowledgements

None.

