## Supplementary File for "Attitudes and Perceptions of Biomedical Journal Editors-in-Chief Towards the Use of Artificial Intelligence Chatbots in the Scholarly Publishing Process: A Cross-Sectional Survey"

### Online Supplement

### eMethods

#### eTable 1: Included Journal Sources

| Publisher | Journal Category | URL |
|---|---|---|
| Elsevier | Medicine and Dentistry | https://www.sciencedirect.com/browse/journals-and-books?contentType=JL&subject=medicine-and-dentistry |
| Elsevier | Nursing and Health Professionals | https://www.sciencedirect.com/browse/journals-and-books?contentType=JL&subject=nursing-and-health-professions |
| Elsevier | Pharmacology Toxicology and Pharmaceutical Science | https://www.sciencedirect.com/browse/journals-and-books?contentType=JL&subject=pharmacology-toxicology-and-pharmaceutical-science |
| Wiley | Dentistry | https://onlinelibrary.wiley.com/action/showPublications?PubType=journal&startPage=&ConceptID=104 |
| Wiley | Health and Healthcare | https://onlinelibrary.wiley.com/action/showPublications?PubType=journal&startPage=&ConceptID=42 |
| Wiley | Medical Science | https://onlinelibrary.wiley.com/action/showPublications?PubType=journal&startPage=&ConceptID=69 |
| Wiley | Nursing and Midwifery | https://onlinelibrary.wiley.com/action/showPublications?PubType=journal&startPage=&ConceptID=78 |
| Springer (part of Springer Nature) | Biomedicine | https://link.springer.com/search?facet-discipline=%22Biomedicine%22&facet-content-type=%22Journal%22 |

| | | |
|---|---|---|
| Springer (part of Springer Nature) | Dentistry | https://link.springer.com/search?facet-content-type=%22Journal%22&facet-discipline=%22Dentistry%22 |
| Springer (part of Springer Nature) | Medicine | https://link.springer.com/search?facet-content-type=%22Journal%22&facet-discipline=%22Medicine%22 |
| Springer (part of Springer Nature) | Medicine and Public Health | https://link.springer.com/search?facet-content-type=%22Journal%22&facet-discipline=%22Medicine+%26+Public+Health%22 |
| Springer (part of Springer Nature) | Pharmacy | https://link.springer.com/search?facet-content-type=%22Journal%22&facet-discipline=%22Pharmacy%22 |
| BMC (Part of Springer Nature) | Biomedicine | https://www.biomedcentral.com/journals#Biomedicine |
| BMC (Part of Springer Nature) | Dentistry | https://www.biomedcentral.com/journals#Dentistry |
| BMC (Part of Springer Nature) | Medicine and Public Health | https://www.biomedcentral.com/journals#Medicine-Public-Health |
| Taylor and Francis | Health and Social Care | https://www.tandfonline.com/topic/allsubjects/hs |
| Taylor and Francis | Medicine, Dentistry, Nursing, and Allied Health | https://www.tandfonline.com/topic/allsubjects/me |
| SAGE | Health Sciences | https://journals.sagepub.com/action/showPublications?category=10.1177/health-sciences |

#### Survey Recruitment Emails

**Initial Recruitment Email**

**Subject Line:** Invitation to Participate in a Survey of Perceptions of Medical Journal Editors Towards the Use of Artificial Intelligence Chatbots (e.g., ChatGPT) in the Scholarly Publishing Process

Dear <NAME>,

I hope this email finds you well. My name is Dr. Jeremy Y. Ng and I am a postdoctoral research associate working under the supervision of Dr. David Moher at the Centre for Journalology at the Ottawa Hospital Research Institute. We obtained a random sample of editor in chiefs (EiCs) of recently publishing biomedical journals. Your name was among those captured in our sample and we are writing to invite you to contribute to a short survey about the use of artificial intelligence (AI) chatbots in the scholarly publishing process.

AI chatbots are programs designed to simulate conversations with human users through text or speech. Some well-known examples include Bing Chat, ChatGPT, YouChat, and Google Bard, although this list is by no means exhaustive. In response to prompts and questions, these AI chatbots can generate relevant responses and material (e.g., text, images, figures, etc.) on a variety of topics. As you may already know, AI chatbots have recently taken the world by storm, and have been used for many purposes, including customer service, mental health support, and education. In recent months, there has been growing interest in the use of AI chatbots in the scholarly editing process, and perhaps you have explored it’s use for the journal editing/publishing process.

This study will be the first to provide a current, large-scale and international perspective on medical EiCs’ perceptions towards the use of AI chatbots in the scholarly publishing process. Your participation will provide valuable insights into how the medical editorial community perceives and accepts the use of AI chatbots in their work. The results of our survey may help inform the development and implementation of policies surrounding the use of AI chatbots in medical publishing process and address potential concerns or barriers that EiCs may have regarding their use.

By completing the survey, you are providing your consent to participate in the study. Your participation is voluntary. Your responses will be completely anonymous. We anticipate the survey taking approximately 15 minutes to complete.

**If you would like to participate in this study, please click on the link below to view the consent form and access the survey: <insert survey link here>**

**Please note that this is an open survey, and thus, we invite you to share the survey link with any medical EiCs within your academic institution and network of collaborators to also participate in our survey.**

We will send all potential participants a reminder email to complete our survey after 1 and 2 weeks from the original invitation, the survey will close after 4 weeks. Please only complete the survey once and kindly ignore the reminders if already completed. Should you have any questions or concerns about our invitation, please do not hesitate to reach out to our principal investigator, Dr. David Moher, PhD at, or me (co-investigator).

Thank you,
Dr. Jeremy Y. Ng, MSc, PhD (on behalf of the research team)

### eResults

#### Demographic Data

##### eTable 2: Medical EICs Demographics, Familiarity, Use and Training on AICs

| **Sex** | **n(%)** |
|---|---|
| Male | 331(67.6) |
| Female | 150(30.6) |
| Prefer not to say | 8(1.6) |
| Prefer to self-describe | 1(0.2) |
| **Age** | |
| 18-24 | 1(0.2) |
| 25-35 | 5(1.02) |
| 36-45 | 47(9.6) |
| 46-55 | 131(26.6) |
| 56-65 | 139(28.3) |
| 65+ | 156(31.7) |
| Prefer not to say | 13(2.6) |
| **Career Stage** | |
| Early career (<5 years) | 5(1.0) |
| Mid-career (5-10 years) | 18(3.7) |
| Senior (>10 years) | 470(95.3) |
| **Current Position** | |
| University/academic faculty | 385(78.3) |
| Academic research staff | 60(12.2) |
| Government | 13(2.6) |
| Clinician | 105(21.3) |
| Pharmaceutical industry/company | 4(0.8) |
| Scholarly communications | 5(1.0) |
| Consultant | 32(6.5) |
| Third sector | 14(2.9) |
| No other roles outside of Editor-in-Chief | 17(3.5) |
| Other | 43(8.7) |
| **Country of Employment (10 Most Popular Shown)** | |
| United States of America | 196(40.3) |
| United Kingdom of Great Britain and Northern Ireland | 48(9.9) |
| Australia | 35(7.2) |
| Canada | 25(5.1) |
| Italy | 21(4.3) |

| | |
|---|---|
| Spain | 17(3.5) |
| France | 16(3.3) |
| Netherlands | 14(2.9) |
| Germany | 13(2.7) |
| India | 10(2.1) |
| **Primary Research Category of Manuscript Submissions to Journal** | |
| Clinical research | 313(63.6) |
| Preclinical research – in vivo | 89(18.1) |
| Preclinical research – in vitro | 66(13.4) |
| Health systems research | 51(10.4) |
| Health services research | 92(18.7) |
| Methods research | 62(12.6) |
| Epidemiological research | 83(16.9) |
| Other | 83(16.9) |
| **Familiarity with AI chatbots** | **n(%)** |
| Very familiar | 64(13.1) |
| Familiar | 325(66.7) |
| Unfamiliar | 67(13.8) |
| Very unfamiliar | 26(5.3) |
| I'm not sure | 5(1.0) |
| **Likelihood of Using AI Chatbots as an EiC in the Future** | |
| Very likely | 30(6.2) |
| Likely | 109(22.4) |
| Unlikely | 123(25.3) |
| Very unlikely | 127(26.1) |
| I am not sure | 98(20.1) |
| **Interested in Learning More/Receiving Training on AI Chatbots Use in Publishing?** | |
| Yes | 302(64.4) |
| No | 55(11.7) |
| Maybe | 112(23.9) |
| **Training Provided by Journal/Publisher on Appropriate AI Tool Use as EiC?** | |
| Yes, and I have taken it | 30(6.2) |
| Yes, but I have not taken it | 47(9.7) |
| No | 262(53.9) |
| I am not sure | 147(30.3) |
| **Has Your Journal or Publisher Implemented Any Policies Surrounding the Use of AI Chatbots in the Scholarly Publishing Process?** | |
| Yes | 240(49.6) |
| No | 106(21.9) |

| | |
|---|---|
| I am not sure | 138(28.5) |
| **How Much Training and Education Do Journal Editors Need to Effectively Use AI Chatbots in Publishing?** | |
| A lot | 211(45.1) |
| Some | 208(44.4) |
| Very little | 16(3.4) |
| None | 6(1.3) |
| I am not sure | 27(5.8) |

#### Supplementary Figures

##### eFigure 1: Biomedical EiCs Use of Various LLMs for Any Use and EiC Specific Use

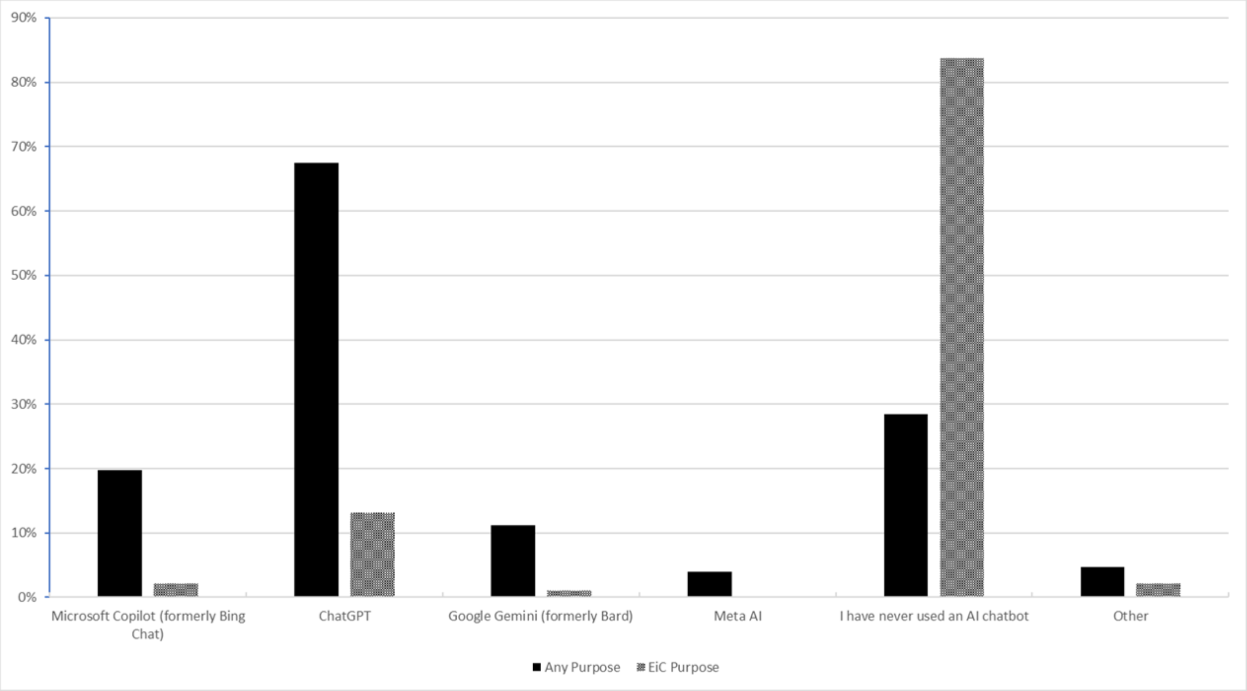

#### eFigure 2: Medical EiCs Perceptions of Potential Future Importance of AICs in Scholarly Publishing and Scientific Research

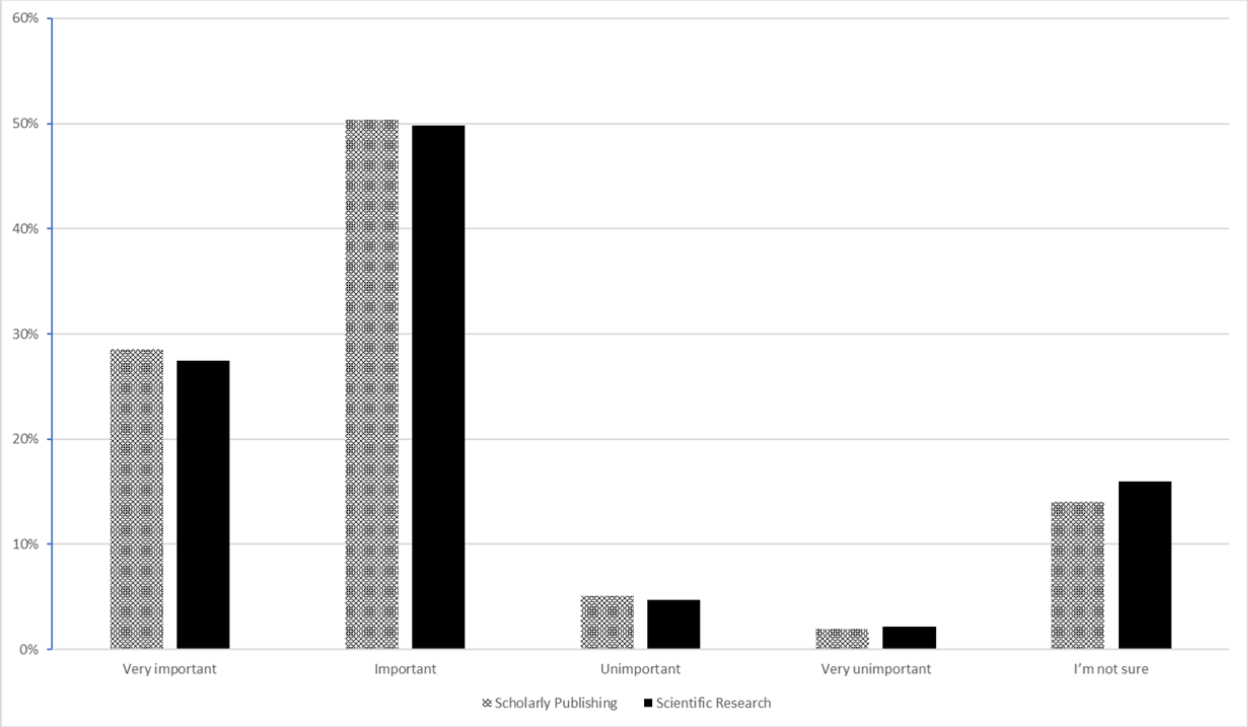

#### eFigure 3. Medical EiCs Perceptions of Potential Future Impact of AICs in Scholarly Publishing and Scientific Research

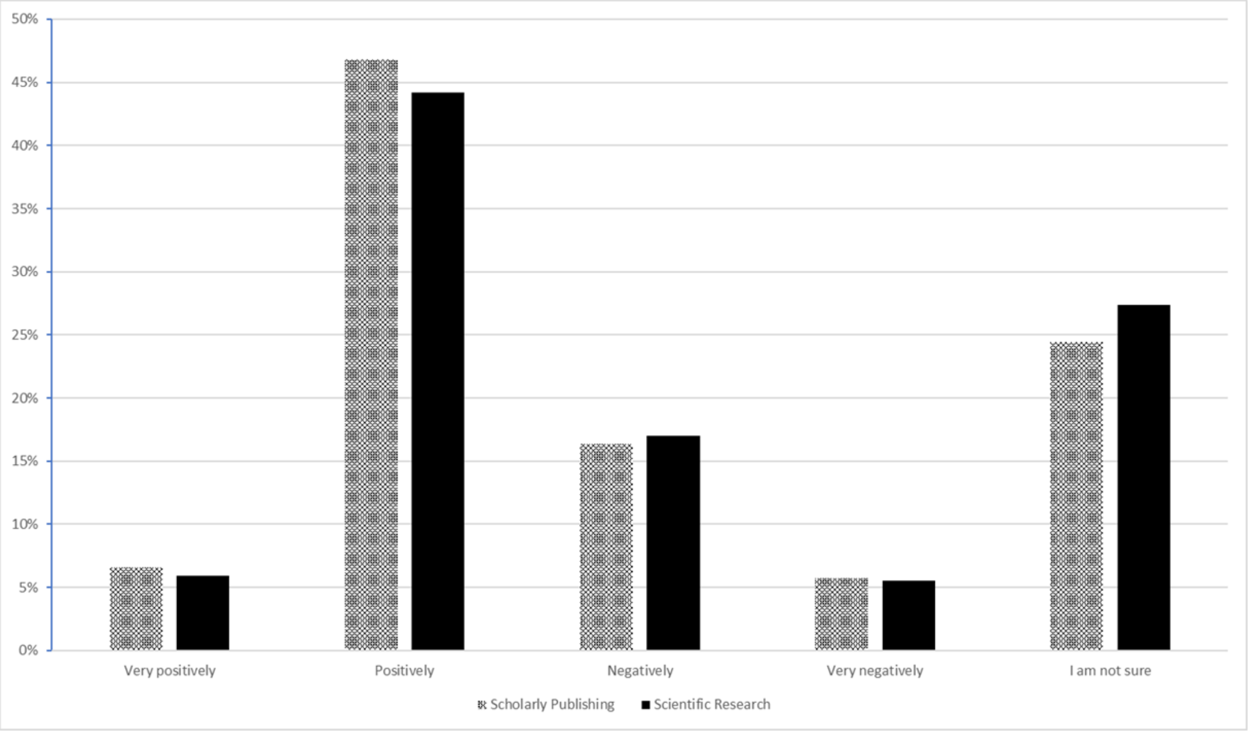

#### Themes from Thematic Analysis of Open-Ended Responses

##### Thematic Analysis eTable 1. Thematic Analysis of the Nature of AI Training Taken by Biomedical Editors-in-Chief (Survey Question 14)

| Themes | Definition and Example |
|---|---|
| **Independent Training** | This theme refers to the Editors in Chief use of independent training to learn about AICs. These include reading publisher policy and guidelines on AICs. |
| **Facilitated Training** | This theme refers to the Editors in Chief use of facilitated training to learn about AICs. These include webinars, seminars and online training. |

##### Thematic Analysis eTable 2. Thematic Analysis of Biomedical Editors-in-Chief Journal/Publisher Policy on AIC Use (Survey Question 16)

| Themes | Definition and Example |
|---|---|

| | |
|---|---|
| **No AI in Authorship or Peer Review** | This theme refers to the Editors in Chief journal/publisher policy on the AIC use, specifically towards no use of AIC in the peer review process as well as inability to list AIC as an author on manuscripts.<br><br>"AI and AI-assisted technologies **should not be listed as authors or co-authors,** nor should they be cited as authors. Authorship carries with it responsibilities and tasks that can only be assigned and performed by humans." |
| **Permitted For Language/Grammar Use** | This theme refers to the permission to use AIC for language and grammar support in Medical Editors in Chief journal/publisher policies on AI use.<br><br>"**The use of generative artificial intelligence (AI)** and AI-assisted technologies by authors in the writing process should be limited to **improving the readability of the text and the language** used" |
| **Need for Declaration of AI Use** | This theme refers to the Editors in Chief journal/publisher policy on the AIC use specifying that all those that use AI need to declare use.<br><br>"**Disclosure by authors is required**. Reviewers are discouraged from chat or use, and disclosure is required." |
| **Confidentiality Concerns with AI in Peer Review** | This theme refers to the journal/publisher policy on AIC use that present confidentiality concerns with the potential use of AI in the peer review process.<br><br>"Editors and peer reviewers **must not upload** files, images or information from unpublished manuscripts **into Generative AI tools**. Failure to comply with this policy may infringe upon the rightsholder's intellectual property." |
| **Other Permissions/Guidelines on AI use** | This theme refers to the broad spectrum of Medical Editors in Chief AIC use journal/publisher policy statements<br><br>"The use of AI is not allowed for the generation of research data, images and figures." |

#### Thematic Analysis eTable 3. Thematic Analysis of Biomedical Editors-in-Chief Perceptions of Steps in the Scholarly Publishing Process wherein AICs Could Be Helpful (Survey Question 18)

| Themes | Definition and Example |
|---|---|
| **Administrative Support** | This term refers to the Medical Editors in Chief perceptions that AiCs could be helpful with general administrative support during the scholarly publishing process.<br><br>**"Writing emails to reviewers and authors."** |
| **Manuscript Preparation** | This term refers to the Medical Editors in Chief perceptions that AiCs could be helpful during the scholarly publishing process.<br><br>**"Writing the manuscript for authors especially if English isn't their first language."** |
| **Supporting Peer Review Process** | This term refers to the Medical Editors in Chief perceptions that AiCs could be helpful with the peer review step of the scholarly publishing process.<br><br>**"**When AI gets extremely good at what it does (maybe 5-10 years from now), I believe **it will be able to serve as an additional peer reviewer for every paper submitted to a journal**, especially if it is given explicit instructions and criteria against which to evaluate a submitted manuscript**."** |
| **Plagiarism and Compliance Assessment** | This term refers to the Medical Editors in Chief perceptions that AiCs could be helpful with plagiarism and compliance assessment during the primary stages of the scholarly publishing process.<br><br>**"**Review of **reporting elements in checklists**, e.g., date of protocol registration, matching of primary outcomes to registration, etc" |

#### Thematic Analysis eTable 4. Thematic Analysis of Steps in the Publishing Process Where Biomedical Editors-in-Chief Have Used AICs for Assistance (Survey Question 20)

| Themes | Definition and Example |
|---|---|

| | |
|---|---|
| **General Administrative Support** | This term refers to the administrative work that the Medical Editors in Chief utilizes AICs for assistance during the scholarly publishing process.<br><br>"I've used AI to develop emails to **communicate with authors**." |
| **Language and Grammar Support** | This term refers to language and grammar supportive work that the Medical Editors in Chief utilizes AICs for assistance during the scholarly publishing process.<br><br>"**Editing my own texts**." |
| **Manuscript Review** | This term refers to the manuscript review step that the Medical Editors in Chief utilizes AICs for assistance during the scholarly publishing process.<br><br>"If the authors use a phrase or refer to some methods I'm not familiar with, I may **ask ChatGPT for a summary** in addition to searching more traditional sources." |

#### Thematic Analysis eTable 5. Thematic Analysis of Biomedical Editors-in-Chief Perceptions of Benefits to the Use of AIC in the Scholarly Publishing Process (Survey Question 28)

| Themes | Definition and Example |
|---|---|
| **Manuscript Quality Assessment** | This term refers to the Medical Editors in Chief perceptions that AICs benefit manuscript quality assessment during the scholarly publishing process.<br><br>"Statistical analysis, reviews, and review of the bibliography are other easy to incorporate roles for chat bots to assist editors" |
| **Language and Grammar Support** | This term refers to the Medical Editors in Chief perceptions that AICs benefit language and grammar during the scholarly publishing process.<br><br>"Mostly in **editing the grammar** from authors in non-English speaking countries." |
| **Administrative Support** | This term refers to the Medical Editors in Chief perceptions that AICs benefits administrative tasks during the scholarly publishing process. |

| | |
|---|---|
| | "Writing responses to inquiries and responding to authors." |
| **General Perceptions of Benefits** | This term refers to the Medical Editors in Chief perceptions that AICs holds no benefit during the scholarly publishing process.<br><br>"None just make the papers worse. Furthermore, they encourage plagiarism." |

#### Thematic Analysis eTable 6. Thematic Analysis of Medical Editors-in-Chiefs Perceptions of Challenges to the Use of AIC in the Scholarly Publishing Process (Survey Question 30)

| **Themes** | **Definition and Example** |
|---|---|
| **Ethical, Integrity, and Privacy Concerns** | This term refers to the Medical Editors in Chief perceptions of ethical, integrity, and privacy challenges with the use of AICs in the scholarly publishing process.<br><br>"The unfamiliarity of Associate Editors of AI is a barrier, particularly when some have been in the profession for a long time. Additionally, there are so many things that can be done with AI, it is hard to keep up, which **may impact the integrity of the journa**l." |
| **Manuscript Quality Concerns** | This term refers to the Medical Editors in Chief perceptions of manuscript quality challenges with the use of AICs in the scholarly publishing process.<br><br>"The biggest threat is the **potential** for authors to use AI chatbots **to fabricate data that cannot be distinguished from real data**." |
| **Negative Impact on Productivity, Publishing Process and Relationships** | This term refers to the Medical Editors in Chief perceptions that challenges with the use of AICs in the scholarly publishing process include a negative impact on the process, relationships, and productivity.<br><br>"**Undermining trust** and interaction dynamics in the editor/reviewer/author relationships." |
| **Integration Challenges** | This term refers to the Medical Editors in Chief perceptions that AIC integration into publishing acts as a challenge with the use of AICs in the scholarly publishing process. |

| | |
|---|---|
| | “The greatest challenge would be **programming the AI chatbox** so it reflects the standards and **guidelines of each editor**. When AI is introduced by a large publishing house, **Chat boxes thus far appear to be too uniform, and not programmed to consider the unique guidelines of each editor**.” |

#### Thematic Analysis eTable 7. Thematic Analysis of Medical Editors in Chief Perceptions of AIC Use in Scholarly Publishing Process (Survey Question 31)

| Themes | Definition and Example |
|---|---|
| **Integration, Ethical, and Confidentiality Implications** | This term refers to the Medical Editors in Chief perceptions of Integration, ethical and confidentiality implications with the use of AICs in the scholarly publishing process.<br><br>**“**As we saw with the Microsoft/Crowdstrike fiasco, technology is great, until it isn't. I **don't believe there are sufficient, reliable systems in place yet to guard against widespread issues.”** |
| **Potential Benefits of AICs** | This term refers to the Medical Editors in Chief perceptions of potential benefits with the use of AICs in the scholarly publishing process.<br><br>**“**The use could **increase productivity** and reduce unnecessary time on many tasks.” |
| **Need for Editor AIC Training** | This term refers to the Medical Editors in Chief perceptions of the need for editor AIC training with the use of AICs in the scholarly publishing process.<br><br>**“EIC need training on AI Chatbot before its real effective use can be implemented effectively in editorial process.** Ineffective and improper use may be counterproductive.” |
| **Study Design Feedback** | This term refers to the Medical Editors in Chief feedback on this study/survey design.<br><br>**“This survey has not touched on the use and abuse of AI by authors and reviewers** which I think is currently the primary concern of editors.” |

| | |
|---|---|
| **Implications on Scholarly Process and Human Aspect** | This term refers to the Medical Editors in Chief perceptions of the implication on scholarly process and human aspect with the use of AICs in the scholarly publishing process.<br><br>**“**I think AI can also be useful in assisting the editorial team but think **we should be much more restrictive as human judgement is essential here**. I am very much against the use of AI in communication with authors and reviewers - even though this takes up a substantial amount of time, **this type of human interaction is essential for a trustworthy peer review and publication process.”** |
| **AIC Limitations** | This term refers to the Medical Editors in Chief perceptions of limitations with the use of AICs in the scholarly publishing process.<br><br>**“**The **check of the quality of fotos submitted** (fotos gross anatomy, fotos of histological specimen) submitted to a morphological journal **does not work by AI alone**.” |
| **Diverse Perspectives on AICs** | This term refers to the Medical Editors in Chief diverse perspectives on AIC integration in the scholarly publishing process.<br><br>“I think a substantial portion of my time as an editors spent in generating communications with authors after synthesizing the input of pure reviewers. This is a low hanging fruit; creating more personalized letters would be extremely helpful and allow me to focus more on strategic goals and troubleshooting Author and publication related issues. **I look forward to learning how CHOP may be able to help with reviewing issues outside the scope of many of my reviewers and myself, including Statistics and methodology. I look forward to being able to use AI in my role as an editor.”** |
